# “Most inactive in my life”: patient-reported barriers to cardiac rehabilitation in heart failure

**DOI:** 10.64898/2026.06.26.26356375

**Authors:** Yulia Khodneva, Megan Nordberg, Todd Brown, Andrea L. Cherrington, Larry Hearld

**Author notes:** **Address for Correspondence:** Yulia Khodneva, MD, PhD 701 19th Street South Birmingham, AL 35294-0007.

## Abstract

**Background & Objective:** Cardiac rehabilitation is an existing guideline-concordant intervention for heart failure that provides benefits but is grossly underutilized by both physicians and patients. We aimed to identify patient-reported barriers and facilitators of participation in cardiac rehabilitation.

**Design, participants, approach:** Qualitative theory-guided in-depth interviews were conducted with adults with heart failure, recruited from ambulatory settings with oversampling of those with heart failure with preserved ejection fraction. Thematic analysis was applied to interview data. Depressive symptoms and perceived stress were assessed by Patient Health Questionnaire (PHQ-8) and Perceived Stress Scale (PSS), respectively.

**Key results:** Twenty-two adults with heart failure, aged 27-85, completed the study; of them 59.1% were women, 68.2% - African American, 4.5% - Hispanic; 77.3% had public insurance or were self-pay; 68.2% had heart failure with preserved ejection fraction. Mean PHQ-8 score was 11.4 (SD= 2.9) and mean PSS score - 20.4 (SD=4.5). Patient-reported barriers to cardiac rehabilitation included unawareness of cardiac rehabilitation and its benefits, perceived inability to exercise, depression, and weight gain, specifically for heart failure with preserved ejection fraction. Perceived inability to exercise stemmed from uncontrolled heart failure symptom burden and exercise intolerance, medication side effects, non-cardiac pain, fear of exercise, and low motivation for exercise. Facilitators of participation included intrinsic and extrinsic motivating factors and specific features of programs, such as individualized and supervised interventions with moderate level of exercise.

**Conclusion:** Participants reported multiple barriers to cardiac rehabilitation; some of them can be modified by providing counselling and referral to cardiac rehabilitation from primary care physicians and simultaneously addressing heart failure symptom burden, pain, stress and depression. Combining cardiac rehabilitation and weight management can benefit adults with heart failure with preserved ejection fraction specifically. Increasing insurance coverage for cardiac rehabilitation for heart failure is warranted.

## Introduction

Heart failure affects over 6 million US adults each year and currently is the only cardiac condition with rising prevalence worldwide^1^. Heart failure patients experience debilitating shortness of breath, increased frailty, reduced functional capacity, markedly decreased quality of life and decreased exercise capacity. Cardiac rehabilitation is an evidence-based multidisciplinary intervention that includes core components of supervised exercise training, lifestyle modifications, and stress management for patients with heart failure^2^. Multiple trials of supervised exercise programs and cardiac rehabilitation have repeatedly demonstrated that exercise is safe for stable patients with heart failure, improves heart failure symptoms and quality of life ^3^, and decreases depression^4^ and mortality.^5^

However, despite the strong evidence that cardiac rehabilitation provides benefit in heart failure, cardiac rehabilitation programs are grossly underutilized in general^6,7^ and among heart failure patients, in particular^8,9^. The data from Get with Guidelines–Heart Failure has shown that of 69,441 patients with heart failure with reduced ejection fraction (HFrEF) who were eligible for cardiac rehabilitation, only 17,076 (24.6%) were referred to cardiac rehabilitation^10^, and only 4.1% of eligible patients with HFrEF participated in the cardiac rehabilitation program.^10^ In the sample restricted to Medicare beneficiaries, enrollment of heart failure patients into cardiac rehabilitation was even lower: only 2.7% of eligible adults with heart failure as their cardiac rehabilitation-qualifying condition used cardiac rehabilitation.^11^

These extremely low rates of enrollment in cardiac rehabilitation suggest that there are important barriers to participation in cardiac rehabilitation, which must be identified to optimize heart failure management. Prior studies found several healthcare system barriers to cardiac rehabilitation utilization among patients with HFrEF, such as poor access to cardiac rehabilitation, stemming from low referral rates from providers, high cost of existing programs, and challenges with transportation to and from the cardiac rehabilitation centers^6^. However, in addition to the system factors, low cardiac rehabilitation utilization can be also explained by a low uptake of the program by the patients with heart failure, with less than 1 out of 20 referred patients finishing the program^10^.

To explore why patients with heart failure do not utilize or complete cardiac rehabilitation programs, we aimed to elicit in-depth patient-reported barriers to exercise and participation in cardiac rehabilitation and identify possible facilitating factors of the participation. We conducted a qualitative study, guided by the “Theory of Planned Behavior”^12^. We oversampled participants with heart failure with preserved ejection fraction (HFpEF) and African American (AA) adults, as these two patient populations are especially affected by the low referral rates and low uptake of cardiac rehabilitation programs. We also compared the barriers to cardiac rehabilitation participation by heart failure subtypes: HFpEF and HFrEF.

## Methods

### Conceptual framework

The semi-structured interview guide created in this study was based on the constructs from the “Theory of Planned Behavior “(TPB). TPB has been used as a conceptual framework guiding studies of the barriers to physical activity among patients with cardiovascular conditions ^13,14^.

The theory includes cognitive constructs, such as behavioral beliefs, attitudes towards behavior, normative beliefs, subjective norms, control beliefs, and perceived behavioral control^15^. These beliefs are thought to influence intention and actual behavior of engaging in exercise^15^. The TPB has been applied to heart failure research, specifically to investigate dietary sodium consumption in heart failure^16^, elicit barriers to exercise in congenital heart disease, and evaluate the feasibility of mHealth application use for home-based cardiac rehabilitation in heart failure.^17^

### Study sample and procedures

Adult English-speaking patients (age ≥ 19 years) diagnosed with stage C heart failure were eligible to participate. Patients with heart failure resulting from congenital heart disease or advanced (stage D) heart failure were excluded because they represented clinically a different patient group, compared to patients with stage C heart failure. Patients with advanced heart failure have more limited functional capacity and much greater safety concerns for performing any physical activity.

Participants were recruited from the primary care and cardiology clinics of an academic health center via direct referral and recruitment. Patients were screened for eligibility using electronic medical records (EMR) and during actual routine clinic visits. Heart failure diagnosis was adjudicated based on manual chart review of physician notes, cardiac imaging, and laboratory data. Individuals, eligible to participate, received a recruitment phone call and were invited to participate in a one-time semi-structured interview.

Sample size determination was guided by thematic saturation and prior studies in methodology of qualitative research which suggested that thematic saturation is usually reached after conducting 15 interviews^18^ and becomes “unwieldy” after data from more interviews are added. We oversampled patients with HFpEF and AA adults with heart failure because they represent a heart failure patient group that is disproportionally affected by worse heart failure outcomes, including worse mortality^1^, is difficult-to-reach, and underrepresented in clinical trials of cardiac rehabilitation interventions^19^. Participants were compensated for their time in a form of a gift card. This study was approved by the Institutional Review Board as an exempt protocol and participants were provided an information sheet prior to interview.

### Data collection

A concurrent mixed methods approach included simultaneous collection of qualitative and quantitative data. Qualitative data were obtained via individual interviews. The interview questions incorporated constructs from the TPB, applied to exercise and heart failure (supplement figure 1) and included items asking participants about current level of physical activity, awareness of cardiac rehabilitation programs, and perceived barriers and facilitators of physical activity. Semi-structured interviews with individual participants were carried out via telephone or a video-platform by a trained research assistant. Interview data were recorded and professionally transcribed. Quantitative data collection included demographics and insurance status, obtained from EMR, interviewer-administered Patient Health Questionaries-8 (PHQ-8), a screening tool for depressive symptoms, and Perceived Stress Scale (PSS) that were collected from the study participants by a research assistant after the qualitative interview was completed.

### Analysis

Qualitative thematic analysis was conducted to identify themes, subthemes, and categories. Three independent coders reviewed and analyzed the interview transcripts separately. Both deductive and inductive analyses were used to identify the themes. For deductive analysis, participants’ responses were coded following the constructs of the TPB. For inductive analysis, coders noted new themes as they arose. Participant responses were separated and contrasted by heart failure subtype: HFrEF and HFpEF. After independent coding, individual coding and analyses of the transcripts were discussed among the coders during coding meetings to create a concise coding tree, eliminate duplicates, and resolve discrepancies in the interpretation of participant responses.

## Results

Between August 2021-April 2022, using outpatient primary care and cardiology clinics’ EMR lists, we identified and contacted 55 eligible patients with heart failure; of them 26 (47.3%) agreed to participate and 22 (40.0%) – completed the interview. Study participants had mean age of 54.0(SD=13.3) years; 68.2% were AA, 4.5% were Hispanic, 59.1% were women, 68.2% had HFpEF (Tables 1 and 2). Forty one percent of the study sample had Medicare or Medicaid public insurance, 36% - were sponsored by the local county healthcare plans with limited coverage for underserved populations and only 23% had private health insurance plans. Mean PHQ8 score was 11.4 (SD= 2.9) and mean PSS score was 20.4 (SD=4.5), indicative of moderate to severe depressive symptoms and high burden of perceived stress, respectively.

**Table 1.** Study Sample.

| Characteristics | N=22 |
| --- | --- |
| Age (years), mean, SD | 54.0 ± 13.3 |
| Race, n (%) |  |
| Black | 15 (68.2) |
| White | 6 (22.7) |
| Hispanic | 1 (4.5) |
| Sex, n (%) |  |
| Female | 13 (59.1) |
| Male | 9 (40.9) |
| Health Insurance, n (%) |  |
| Private plans | 5 (22.7) |
| Medicare/Medicaid | 9 (40.9) |
| County plan (limited coverage) | 8 (36.4) |
| Subtype of heart failure, n (%) |  |
| HFrEF | 7 (31.8) |
| HFpEF | 15 (68.2) |
| PHQ-8, mean, SD | 11.4 ± 2.9 |
| Perceived stress scale, mean, SD | 20.4 ± 4.5 |

Abbreviations: BCBS- Blue Cross/Blue Shield. heart failure- heart failure, HFrEF- heart failure with reduced ejection fraction, HFpEF – heart failure with preserved ejection fraction, SD – standard deviation. \*"County plan" is a plan sponsored by the county department of health and providing limited coverage of health services for individuals without commercial insurance and income below the federal poverty line.

**Table 2.**
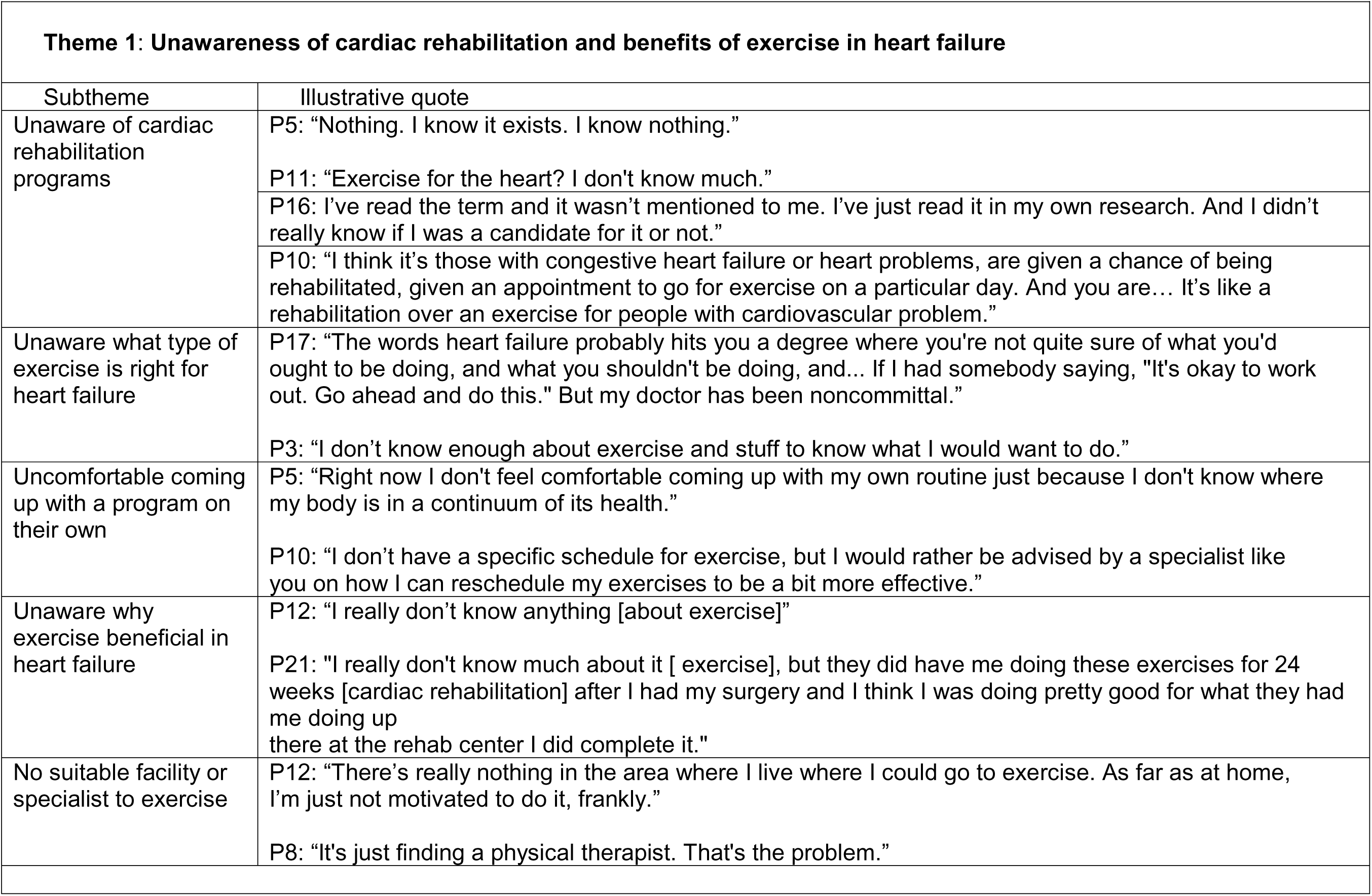
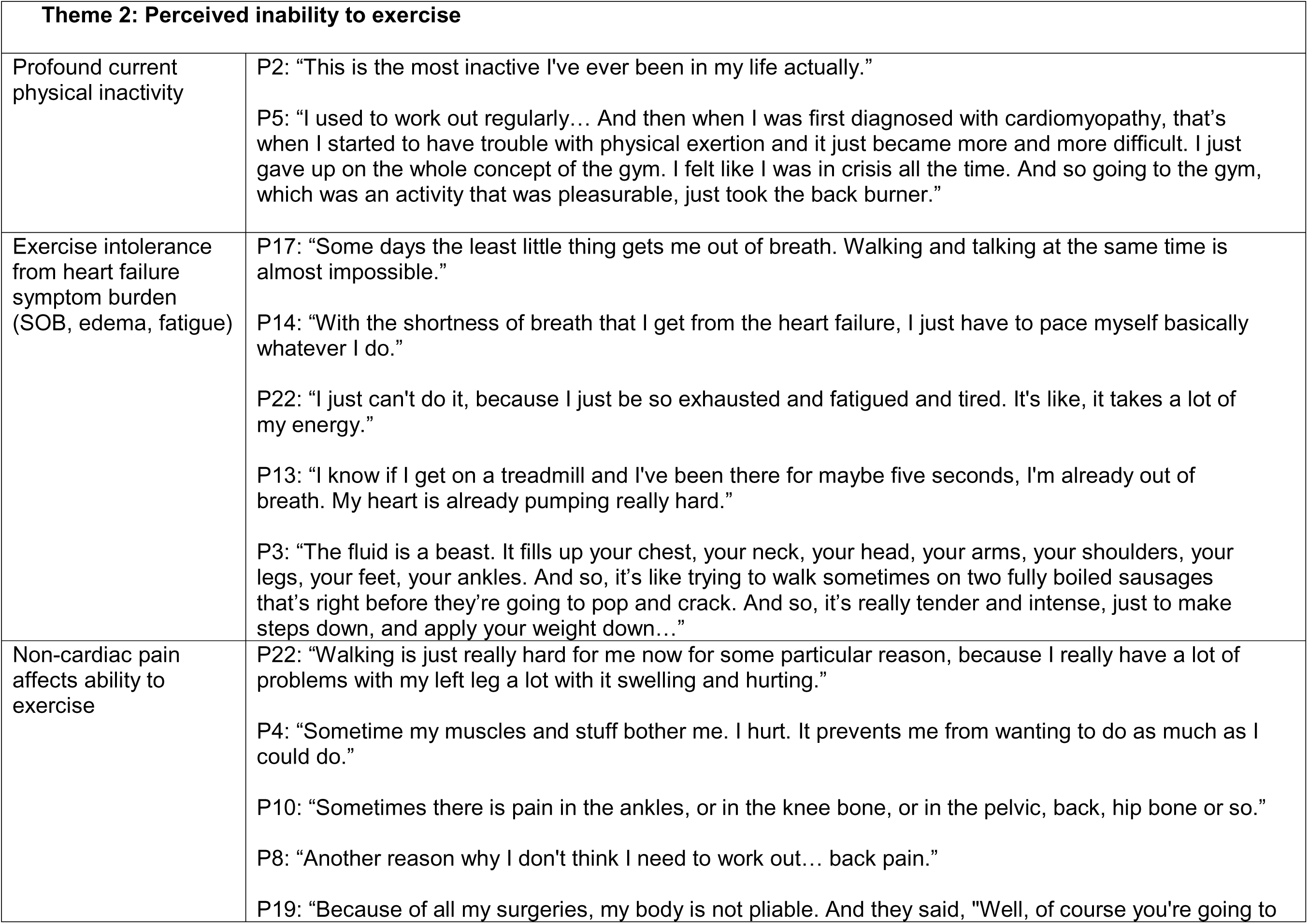
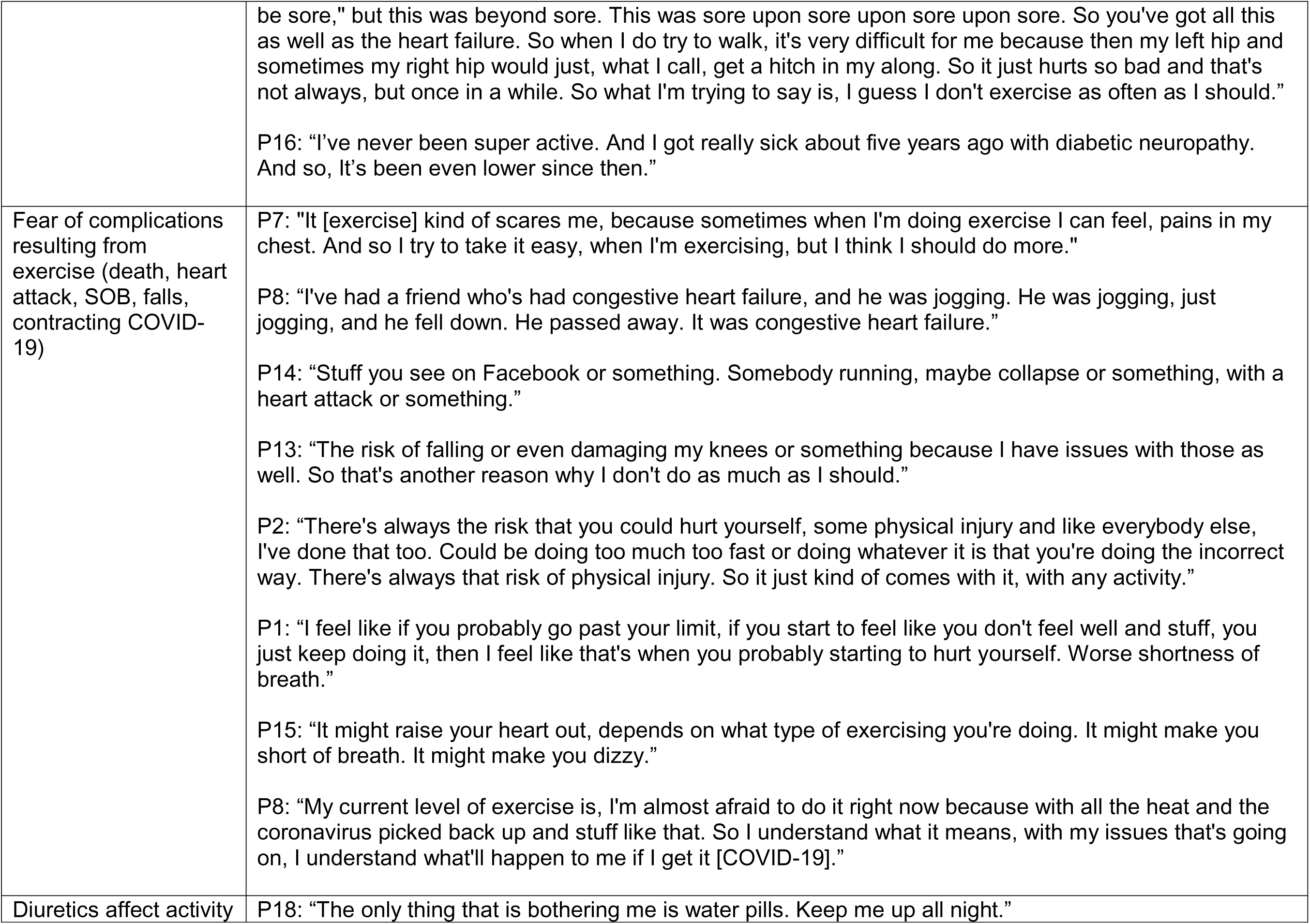
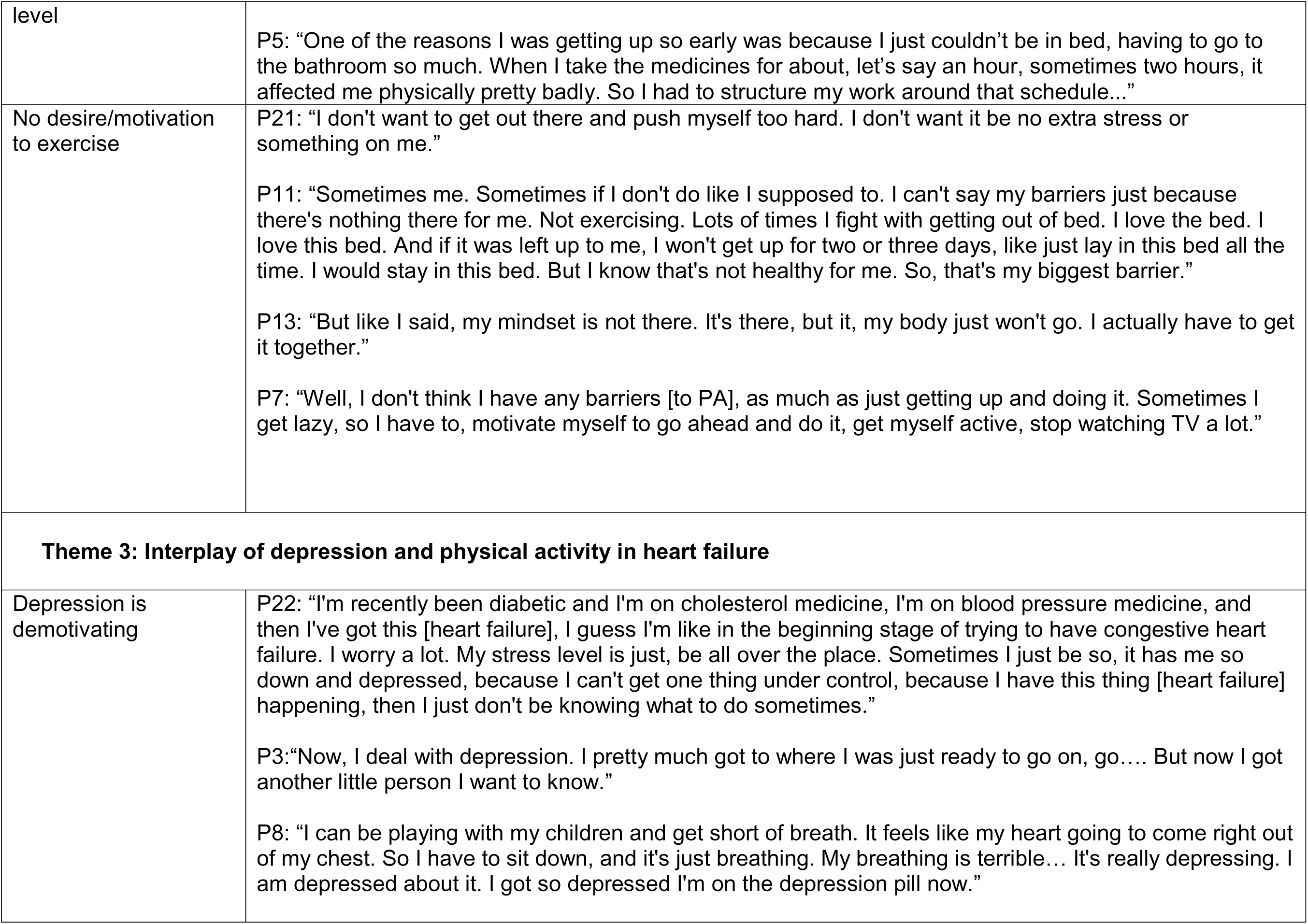
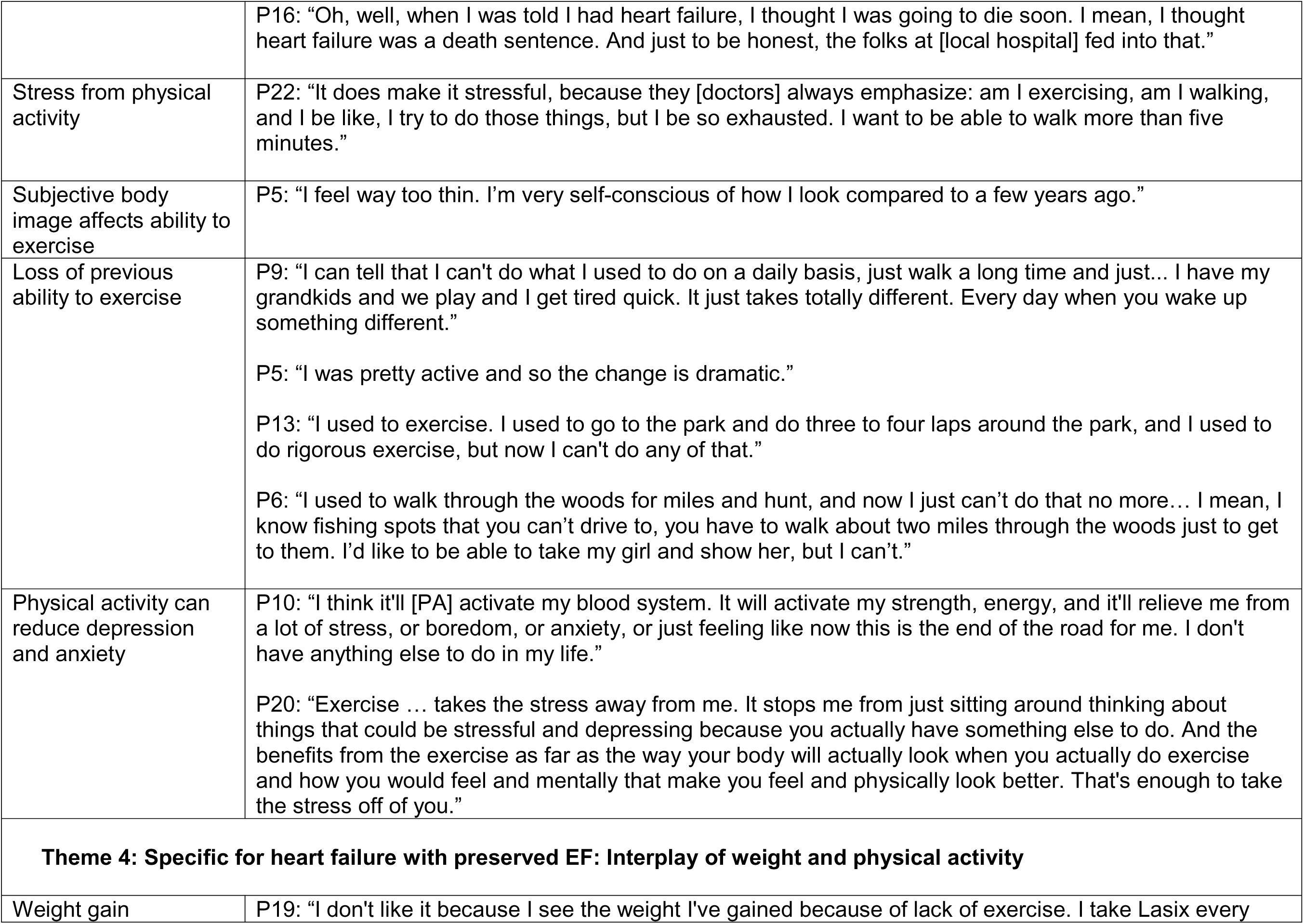
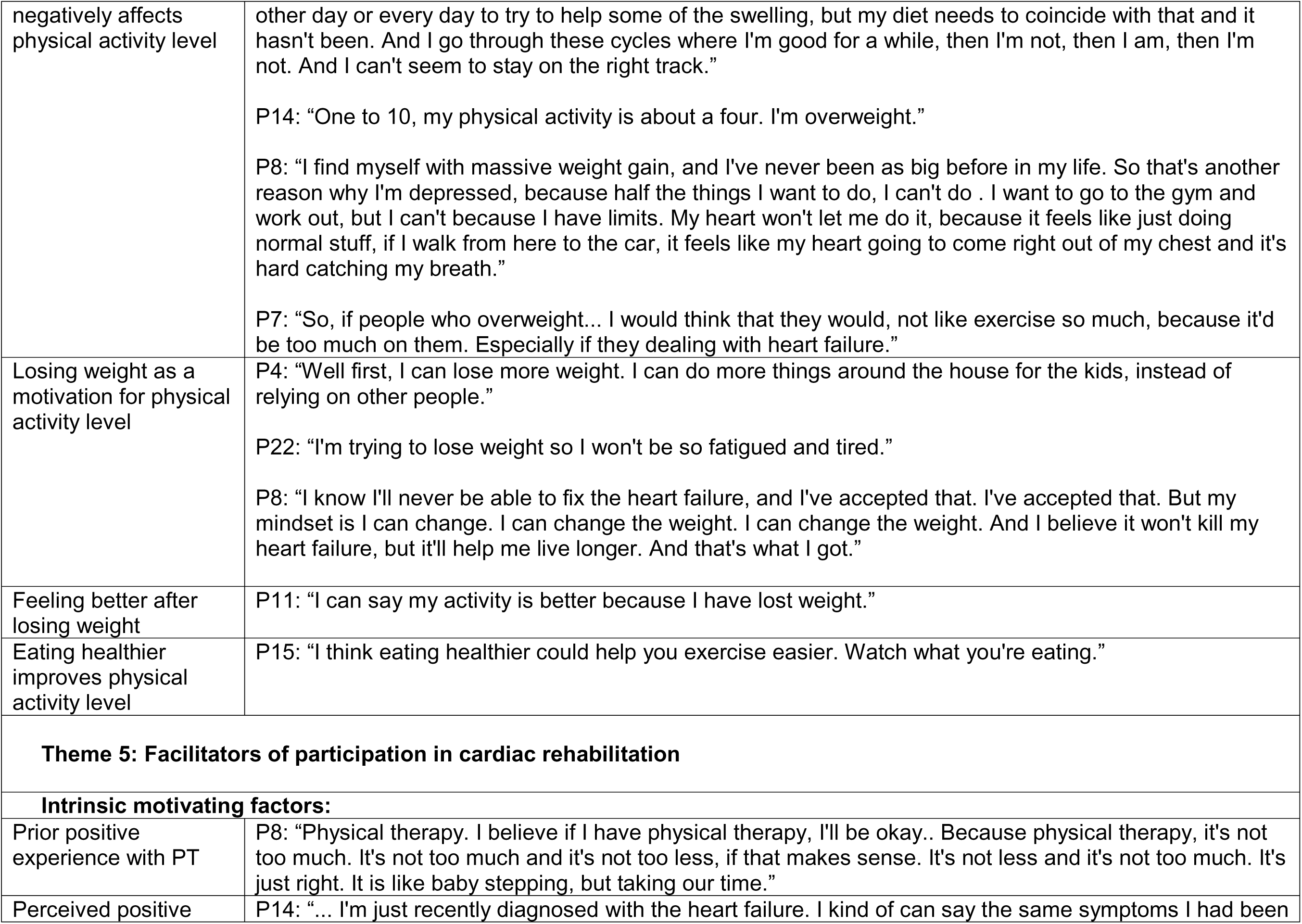
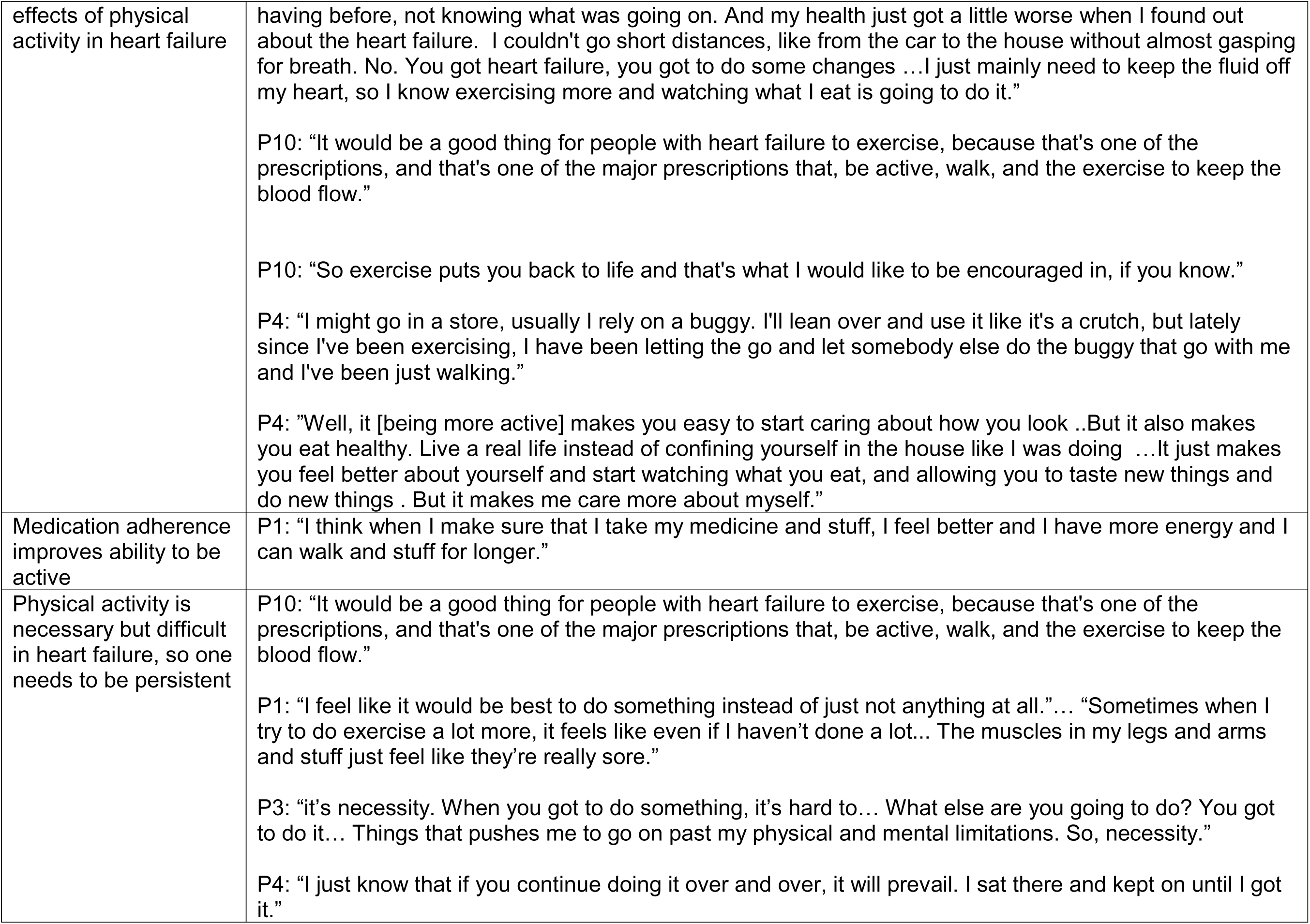
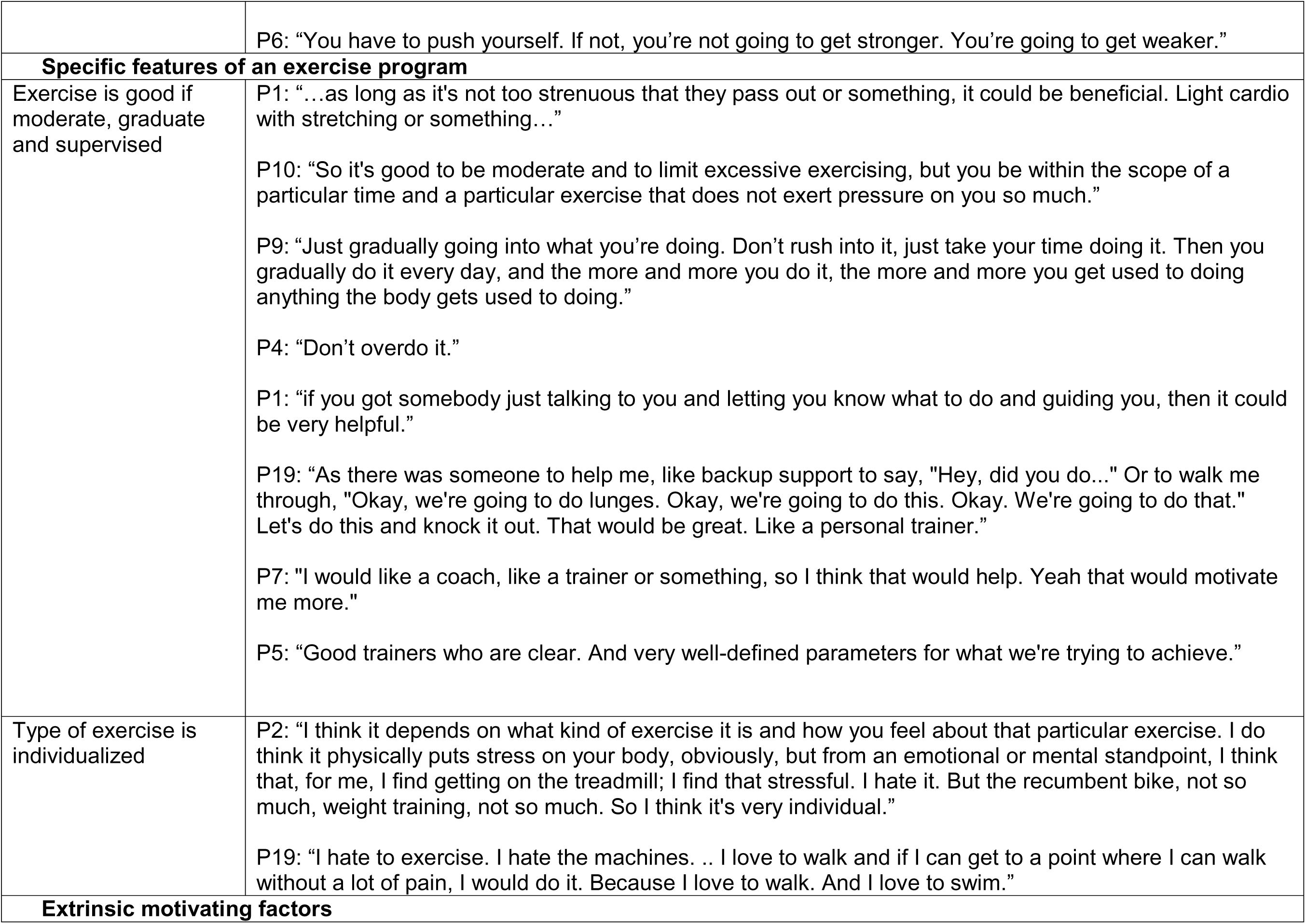
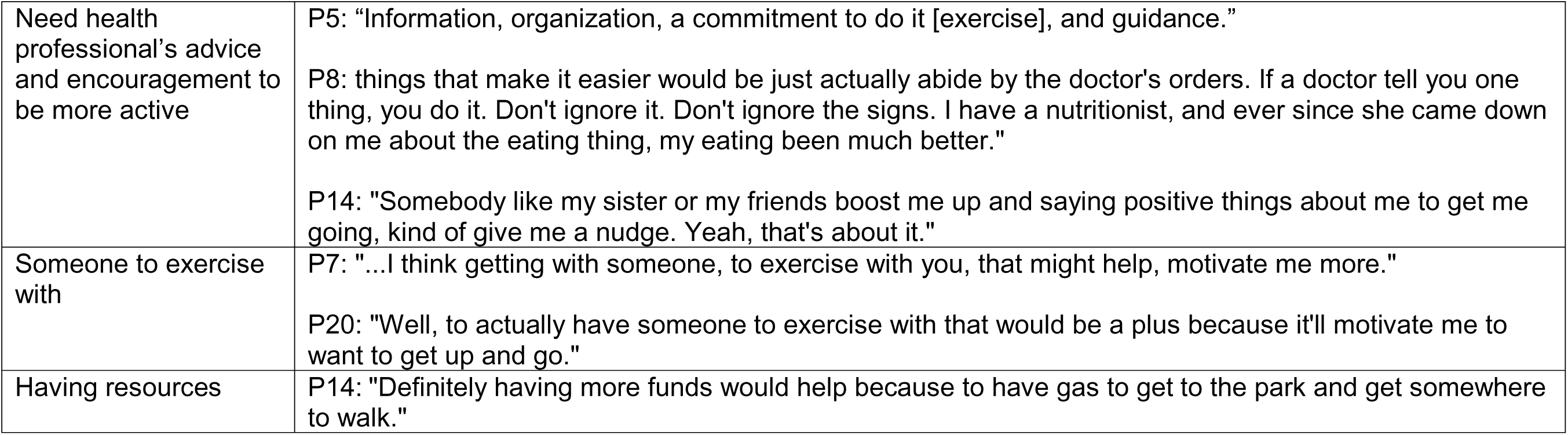
Summary of themes and subthemes.

Qualitative interviews were 20-30 minutes long. Overall, most participants did not know what cardiac rehabilitation programs were: of 22 study participants, only 2 participants underwent cardiac rehabilitation for non-heart failure qualifying conditions and 2 other participants did physical therapy in the past for non-cardiac conditions.

### Thematic analysis

We achieved thematic saturation on the 22^nd^ interview, and therefore data collection was stopped after the 22^nd^ interview. As a result of thematic analysis, we identified 5 themes: unawareness of cardiac rehabilitation programs and their benefits, perceived inability to exercise, interplay of depression and physical activity, interplay of weight and physical activity, and facilitators of participation in cardiac rehabilitation programs (Figure 1 and 2, Table 2).

**Figure 1.**
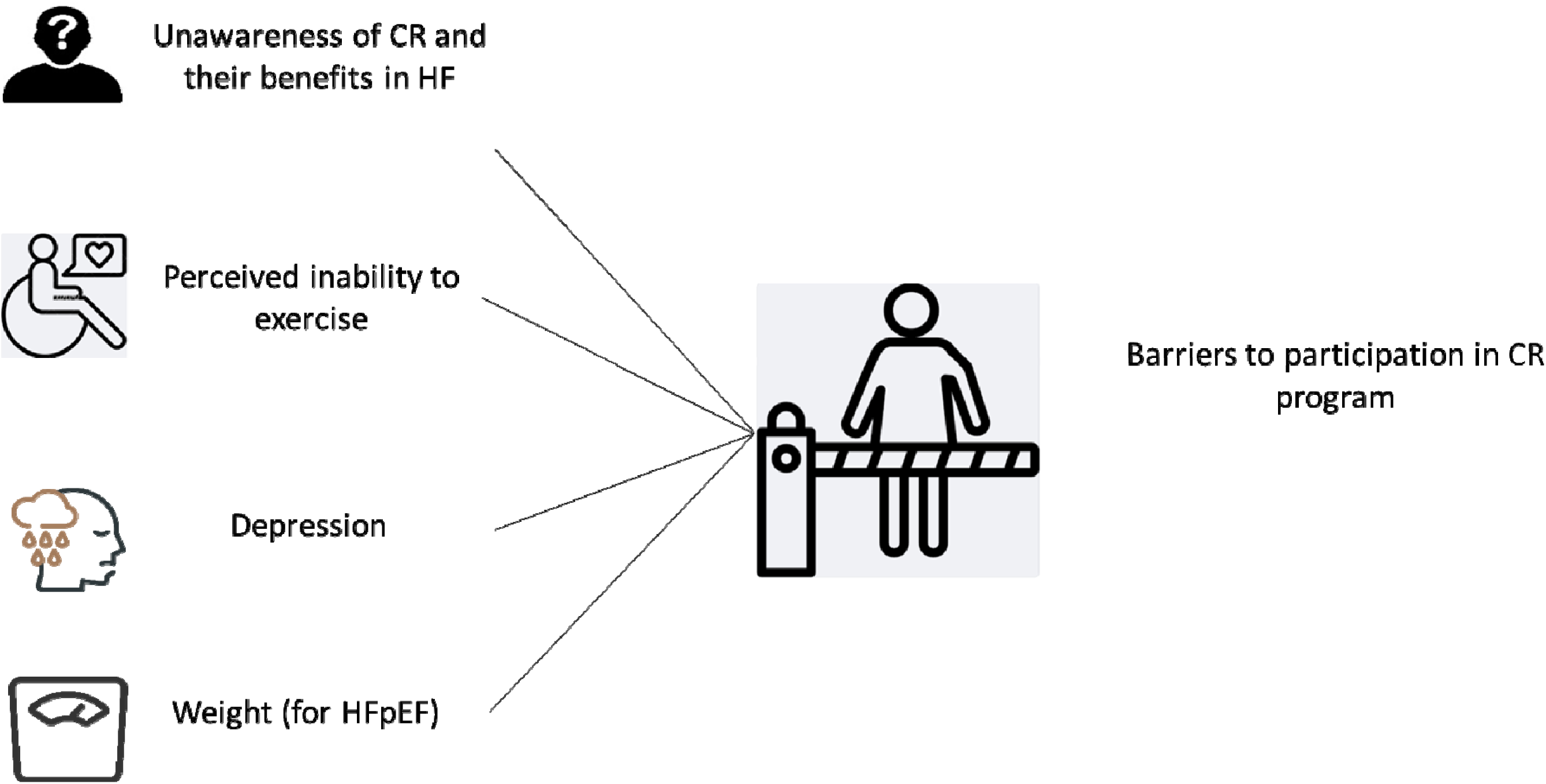
Patient-reported barriers to participation in cardiac rehabilitation in heart failure. Figure 1 Legend: Figure 1 presents 4 themes reflecting barriers to participation in cardiac rehabilitation (CR) reported by participants with heart failure (HF).

**Figure 2.**
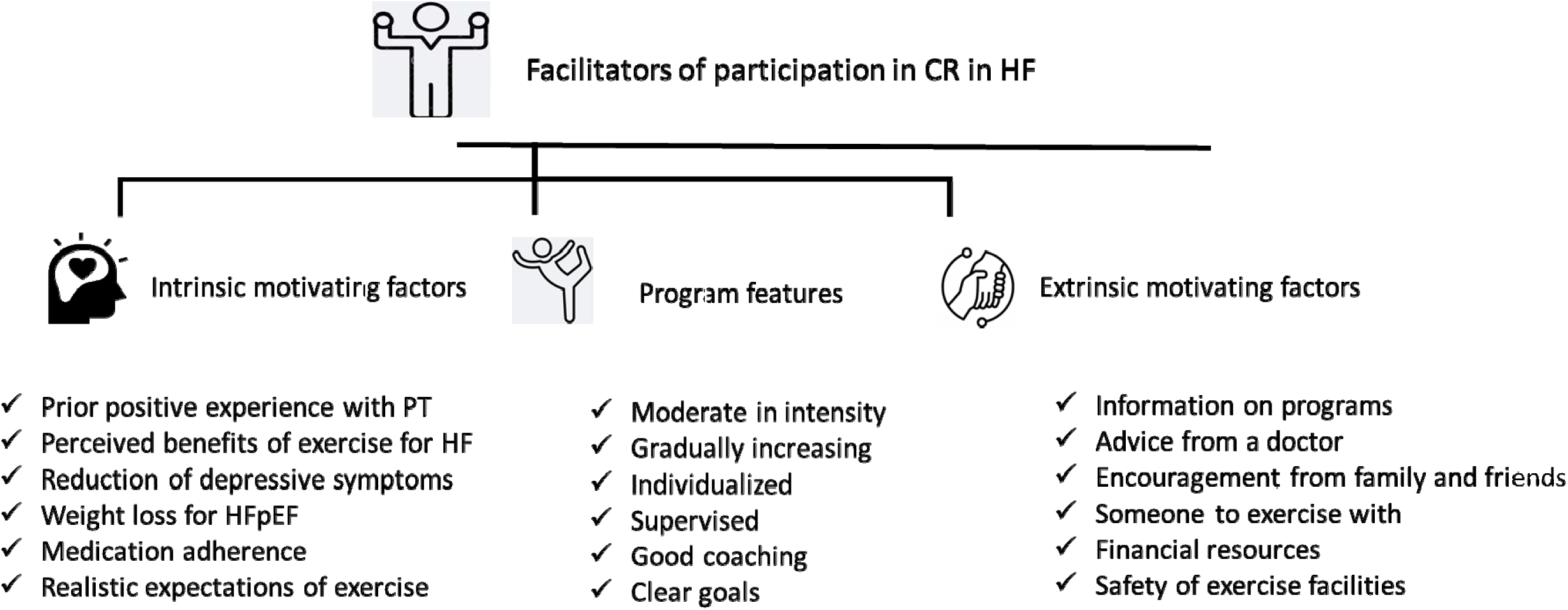
Patient-reported facilitators of participation in cardiac rehabilitation programs. Figure 2 Legend: Figure 2 presents 3 themes, reflecting facilitators of participation in cardiac rehabilitation (CR) or supervised exercise program as reported by participants with heart failure (HF).

When we compared participant responses by the two major subtypes of heart failure, the themes and subthemes were similar between HFrEF and HFpEF, except the theme on the role of weight in physical activity was only reported by participants with HFpEF.

### Theme 1: Unawareness of cardiac rehabilitation programs and their benefits

Most of our study participants noted that they were not aware of cardiac rehabilitation for heart failure. As one participant reported: “Exercise for the heart? I don’t know much” (P11, female with HFpEF). Only one participant attended a cardiac rehabilitation program, but for indications other than heart failure. Some participants read about a cardiac rehabilitation program on their own, as one participant commented: “I’ve read the term but it wasn’t mentioned to me. I’ve just read it in my own research. And I didn’t really know if I was a candidate for it or not” (P16, female with HFpEF). Even if participants reported they heard about cardiac rehabilitation, they did not know the specifics: “Nothing. I know it [cardiac rehabilitation] exists. I know nothing” (P5, male with HFrEF).

Study participants reported that they were unaware of the general benefits of exercise in heart failure: “I really don’t know anything [about exercise]” (P12, female with HFrEF). Participants reported that they did not know what specific type of physical activity is indicated and beneficial for heart failure specifically. As one participant mentioned: “The words heart failure probably hits you a degree where you’re not quite sure of what you’d ought to be doing, and what you shouldn’t be doing, and… If I had somebody saying, ‘It’s okay to work out. Go ahead and do this.’ But my doctor has been noncommittal” (P17, female with HFpEF). Consequently, participants were not comfortable with creating their own exercise program: “Right now I don’t feel comfortable coming up with my own [exercise] routine just because I don’t know where my body is in a continuum of its health” (P5, male with HFrEF). Participants required more guidance on exercise from their healthcare team: “I don’t have a specific schedule for exercise, but I would rather be advised by a specialist like you on how I can reschedule my exercises to be a bit more effective” (P10, male with HFrEF).

### Theme 2: Perceived inability to exercise

Most participants in our study reported profound physical inactivity, as one participant stated, “This is the most inactive I’ve ever been in my life actually” (P2, female with HFpEF). As a result, most of the participants questioned their ability to engage in an exercise program, as one participant shared: “I just can’t do it [exercise], because I just be so exhausted and fatigued and tired. It’s like, it takes a lot of my energy” (P22, female with HFpEF).

Participants reported several factors contributed to their physical inactivity. One of the most significant barriers was exercise intolerance from heart failure symptoms, such as shortness of breath, lower extremity edema, and fatigue. As one participant stated, “With the shortness of breath that I get from the heart failure, I just have to pace myself basically whatever I do” (P14, male with HFpEF). And another participant shared: “The fluid is a beast. It fills up your chest, your neck, your head, your arms, your shoulders, your legs, your feet, your ankles. And so, it’s like trying to walk sometimes on two fully boiled sausages that’s right before they’re going to pop and crack. And so, it’s really tender and intense, just to make steps down, and apply your weight down…” (P3, female with HFrEF).

Even those individuals who had a prior active lifestyle reported dramatically reduced ability to exercise after heart failure diagnosis: “I used to work out regularly… And then when I was first diagnosed with cardiomyopathy, that’s when I started to have trouble with physical exertion and it just became more and more difficult. I just gave up on the whole concept of the gym. I felt like I was in crisis all the time. And so going to the gym, which was an activity that was pleasurable, just took the back burner” (P5, male with HFrEF).

Study participants also reported on the negative effects of heart failure medications on their physical activity level. Specifically, they mentioned that diuretics negatively affected sleep and were causing daytime sleepiness so they could not exercise during the day: “The only thing that is bothering me is water pills. Keep me up all night” (P18, male with HFpEF). Another problem reported by participants related to diuretics was that their effect was disruptive for their routine. “When I take the medicines for about, let’s say an hour, sometimes two hours, it affected me physically pretty badly. So I had to structure my work around that schedule.” (P5, male with HFrEF).

Another contributing factor to participants’ perceived inability to exercise was a high level of chronic non-cardiac pain. As one patient shared, “Because of all my surgeries [for arthritis], my body is not pliable… So you’ve got all this as well as the heart failure. So when I do try to walk, it’s very difficult for me because then my left hip and sometimes my right hip would just, what I call, get a hitch in my along. So it just hurts so bad and that’s not always, but once in a while. So what I’m trying to say is, I guess I don’t exercise as often as I should” (P19, female with HFpEF).

Furthermore, a very important contributor to perceived inability to exercise was patients’ fear of exercise and, specifically, fear of heart failure complications from increased physical activity.

One participant reported on the death of a friend while jogging: “I’ve had a friend who’s had congestive heart failure, and he was jogging. He was jogging, just jogging, and he fell down. He passed away. It was congestive heart failure” (P8, male with HFpEF). Some participants reported that fear of exercise in heart failure was supported by stories that they read on social media when physical activity resulted in bad outcomes for heart failure patients: “Stuff you see on Facebook or something. Somebody running, maybe collapse or something, with a heart attack or something” (P14, male with HFpEF).

For some study participants, fear of exercise was more related to mechanical falls during exercise and a risk of contracting COVID19. As one participant stated: “The risk of falling or even damaging my knees or something because I have issues with those as well. So that’s another reason why I don’t do as much as I should” (P13, female with HFpEF). Since the data collection predominantly occurred during the Covid-19 pandemic, participants also reflected on fear of contracting COVID in shared exercise facilities: “My current level of exercise is, I’m almost afraid to do it right now because with all the heat and the coronavirus picked back up and stuff like that. So I understand what it means, with my issues that’s going on, I understand what’ll happen to me if I get it [COVID-19]” (P8, male with HFpEF).

Finally, some participants shared that they had no desire to exercise: “I don’t want to get out there and push myself too hard. I don’t want it be no extra stress or something on me” (P21, male with HFpEF).

### Theme 3: Interplay of depression and exercise in heart failure

Analysis of the PHQ8 and PSS scales demonstrated a high level of both depressive symptoms and perceived stress among our study participants with heart failure. Matching our quantitative findings, during interviews, participants with heart failure shared high levels of stress and depression in their lived experiences. According to participants’ reports, the relationships between depression and heart failure were complex and bi-directional. For some participants, depressive symptoms and high stress levels decreased their motivation to be active. For another group of participants, engaging in any amount of exercise reduces depression and perceived stress. For example, as one participant reflected on stress and depression in their life: “I’m recently been diabetic and I’m on cholesterol medicine, I’m on blood pressure medicine, and then I’ve got this [heart failure], I guess I’m like in the beginning stage of trying to have congestive heart failure. I worry a lot. My stress level is just, be all over the place. Sometimes I just be so, it has me so down and depressed, because I can’t get one thing under control, because I have this thing [heart failure] happening, then I just don’t be knowing what to do sometimes” (P22, female with HFpEF). Depressive symptoms stemmed from uncontrolled heart failure symptoms, as one participant explained: “I can be playing with my children and get short of breath. It feels like my heart going to come right out of my chest. So I have to sit down, and it’s just breathing. My breathing is terrible… It’s really depressing. I am depressed about it. I got so depressed I’m on the depression pill now” (P8, male with HFpEF).

Participants also reflected that needing to be active while living with heart failure was stressful for them, as one participant pointed out: “It [A need to exercise] does make it stressful, because they [doctors] always emphasize: am I exercising, am I walking, and I be like, I try to do those things, but I be so exhausted. I want to be able to walk more than five minutes” (P22, female with heart failure).

Participants with heart failure were also self-conscious about their body image and reflected how it affected their motivation to visit exercise facilities: “I feel way too thin. I’m very self-conscious of how I look compared to a few years ago” (P5, male with HFrEF).

A large group of study participants expressed a loss of previous ability to be active and to exercise after heart failure diagnosis. As one participant stated “I used to exercise. I used to go to the park and do three to four laps around the park, and I used to do rigorous exercise, but now I can’t do any of that” (P13, female with HFpEF).Participants reported how development of heart failure caused them to drop their favorite activities such as hiking, hunting, and fishing and caused them to experience negative emotions: “I used to walk through the woods for miles and hunt, and now I just can’t do that no more… I mean, I know fishing spots that you can’t drive to, you have to walk about two miles through the woods just to get to them. I’d like to be able to take my girl and show her, but I can’t” (P6, male with HFrEF).

On the contrary, for a smaller group of our study participants, even a small amount of physical activity has produced positive effects and reduced their level of depression and anxiety: “Exercise … takes the stress away from me. It stops me from just sitting around thinking about things that could be stressful and depressing because you have something else to do. And the benefits from exercise as far as the way your body will actually look when you actually do exercise and how you would feel and mentally that make you feel and physically look better. That’s enough to take the stress off of you” (P20, female with HFpEF).

### Theme 4: Specific for HFpEF: Interplay of weight and physical activity

Compared to participants with HFrEF, participants with HFpEF specifically reported how their weight and physical activity were interrelated. For some participants with HFpEF, recent weight gain impeded their physical activity. As one participant stated, “I find myself with massive weight gain, and I’ve never been as big before in my life. …half the things I want to do, I can’t do. I want to go to the gym and work out, but I can’t because I have limits” (P8, female with HFpEF). One participant also shared a concern that if heart failure patients are overweight, it would be too much for them to exercise in general: “So, if people who overweight… I would think that they would, not like exercise so much, because it’d be too much on them. Especially if they are dealing with heart failure” (P7, male with HFpEF).

Participants reflected on the struggle between attempts to lose weight and their ability to adhere to healthy diet and exercise schedule: “I don’t like it because I see the weight I’ve gained because of lack of exercise. I take Lasix every other day or every day to try to help some of the swelling, but my diet needs to coincide with that and it hasn’t been. And I go through these cycles where I’m good for a while, then I’m not, then I am, then I’m not. And I can’t seem to stay on the right track” (P19, female with HFpEF). Some participants also reflected on how being overweight produces physiological changes and symptoms such as worsening shortness of breath and palpitations.

For another group of study participants, a weight loss goal was a motivator for physical activity. Specifically, they mentioned that if they lost more weight, they were able to do more around the house and for the kids: “Well first, I can lose more weight. I can do more things around the house for the kids, instead of relying on other people” (P4, female with HFpEF). They were hopeful that losing weight would promote a decrease in acuity of heart failure symptoms: “I’m trying to lose weight so I won’t be so fatigued and tired” (P22, female with HFpEF). They also shared a deep understanding of heart failure as a disease and said that even if they could not reverse heart failure completely, they could contribute to losing weight and adopting healthier lifestyles: “I know I’ll never be able to fix the heart failure, and I’ve accepted that. I’ve accepted that. But my mindset is I can change. I can change the weight. I can change the weight. And I believe it won’t kill my heart failure, but it’ll help me live longer. And that’s what I got” (P8, female with HFpEF).

Those participants with HFpEF who were able to achieve their desired weight loss shared that their ability to exercise had already improved: “I can say my activity is better because I have lost weight” (P11, female with HFpEF). Finally, participants mentioned that adherence to a healthy diet contributed to better exercise levels: “I think eating healthier could help you exercise easier. Watch what you’re eating” (P15, female with HFpEF).

### Theme 5: Facilitators of participation in cardiac rehabilitation

Despite many study participants reporting on a very inactive lifestyle, most of them were interested in finding ways to be more active even while living with heart failure. They shared a belief that they could engage in physical activity and cardiac rehabilitation programs if the programs were presented to them clearly and organized in a person-centered way. Facilitators of exercise included intrinsic motivating factors, specific features of an exercise program, and extrinsic motivating factors (Figure 2).

One of the most important intrinsic motivating factors for being more active was a previous positive experience with physical activity and supervised exercise program in the form of physical therapy. One participant shared their experiences with being involved in a physical therapy program: “Physical therapy. I believe if I have physical therapy, I’ll be okay… Because physical therapy, it’s not too much. It’s not too much and it’s not too less, if that makes sense. …It’s just right. It is like baby stepping, but taking our time” (P8, male with HFpEF).

Even without prior positive experience with cardiac rehabilitation programs, study participants theorized about potential positive effects of being more physically active. One participant shared that exercise may help reduce heart failure symptoms: “I just mainly need to keep the fluid off my heart, so I know exercising more and watching what I eat is going to do it” (P14, male with HFpEF). Another participant reported that physical activity helped them be more social: “Live a real life instead of confining yourself in the house like I was doing… It just makes you feel better about yourself and start watching what you eat, and allowing you to taste new things and do new things” (P4, female with HFpEF).

Better control over heart failure symptoms, specifically through medication adherence, was another intrinsic motivating factor to improve ability to exercise. As one participant stated, “I think when I make sure that I take my medicine and stuff, I feel better and I have more energy and I can walk and stuff for longer” (P1, female with HFrEF).

Finally, a realistic expectation that exercise is difficult but necessary in heart failure was a facilitator of adherence to exercise programs. As one of the participants noted, “You have to push yourself. If not, you’re not going to get stronger. You’re going to get weaker” (P6, male with HFrEF).

Most participants reported that they wanted moderate intensity exercise along with good supervision and clear instructions during a supervised exercise program. As one participant stated, “So it’s good to be moderate and to limit excessive exercising, but you be within the scope of a particular time and a particular exercise that does not exert pressure on you so much” (P10, male with HFrEF). Supervision during exercise was extremely important for our study participants: “If you got somebody just talking to you and letting you know what to do and guiding you, then it could be very helpful” (P1, female with HFrEF).

Additionally, participants emphasized that the cardiac rehabilitation program should be individualized according to patient needs and preferences. One participant stated, “I think it depends on what kind of exercise it is and how you feel about that particular exercise. I do think it physically puts stress on your body, obviously, but from an emotional or mental standpoint, I think that, for me, I find getting on the treadmill; I find that stressful. I hate it. But the recumbent bike, not so much, weight training, not so much. So I think it’s very individual” (P2, female with HFpEF).

Extrinsic motivation for engaging in cardiac rehabilitation programs, as emphasized by our study participants, was the need to be prepared for such a program and to have more information: “Information, organization, a commitment to do it [exercise], and guidance” (P5, male with HFrEF).

Study participants reported that to engage in a supervised exercise program they would need a recommendation from their doctor or another health care professional: “Things that make it [cardiac rehabilitation program participation] easier would be just actually abide by the doctor’s orders. If a doctor tell you one thing, you do it. Don’t ignore it. Don’t ignore the signs. I have a nutritionist, and ever since she came down on me about the eating thing, my eating been much better” (P8, male with HFpEF).

Another important extrinsic motivating factor was receiving encouragement from their social network to be more active. One participant explained, “Somebody like my sister or my friends boost me up and saying positive things about me to get me going, kind of give me a nudge. Yeah, that’s about it” (P14, male with HFpEF). Some participants mentioned that they would prefer to attend exercise programs with a friend: “Well, to actually have someone to exercise with that would be a plus because it’ll motivate me to want to get up and go” (P20, female with HFpEF).

Finally, our participants underscored a need to have adequate resources to afford physical therapy or exercise program copays, gas and transportation to facilities, and safety of public outside exercise facilities. As, for example, one of the participants noted: “Definitely having more funds would help because to have gas to get to the park and get somewhere to walk” (P14, male with HFpEF).

## Discussion

Most of our study participants with heart failure were not referred to cardiac rehabilitation and did not have direct experience with cardiac rehabilitation programs. This study identified several patient-reported barriers to participation in cardiac rehabilitation programs. Barriers to cardiac rehabilitation participation, relevant to both participants with HFpEF and participants with HFrEF, were unawareness that cardiac rehabilitation programs exist and are beneficial, perceived inability to exercise, and depression. Specifically, perceived inability to exercise was closely related to a very low level of current physical activity among our study sample, stemming from uncontrolled heart failure burden, diuretic use side effects, fear of exercise, non-cardiac chronic pain and low motivation. Being overweight was a specific barrier reported by participants with HFpEF only.

At the same time, study participants offered their suggestions on factors that can boost their participation in cardiac rehabilitation programs, summarized into intrinsic and extrinsic motivation and specific features of the program. Previous positive experiences with physical therapy and perceived benefits of exercise, such as improvement of heart failure symptoms, being more independent, increased ability to socialize, and feeling good about one’s body were among the intrinsic motivating factors. For another group of our study participants, extrinsic motivating factors such as encouragement from family, having an exercise partner, safety of the exercise facilities, and advice from their doctor, were important.

The 2022 American Heart Association (AHA)/American College of Cardiology (ACC) guidelines for heart failure management included cardiac rehabilitation into a class IA recommendation for heart failure patients^20,21^ and specified that cardiac rehabilitation can be useful in improving functional capacity, exercise tolerance, and health-related quality of life for patients with heart failure (class IIA recommendation). To follow this recommendation and to illicit patient perspective, we investigated what features of cardiac rehabilitation programs are attractive to individuals with heart failure. Participants shared that they are more likely to participate in a cardiac rehabilitation program that is individualized and moderately intense, with a gradual increase in intensity. They also emphasized a need for supervision and having a good coach who will set clear goals. Most of existing cardiac rehabilitation programs already include all the features desired by heart failure patients, but unfortunately, as our study showed, most heart failure patients are not informed about these programs and are not counselled by their healthcare teams about the benefits of exercise in heart failure. Specifically, in our study sample, most of the participants had Medicaid/Medicare insurance or limited county-sponsored health plans that had limited coverage of cardiac rehabilitation referrals. Limited health insurance coverage could explain a very low level of knowledge of cardiac rehabilitation programs among participants with heart failure in our study.

To our knowledge, this is one of the first studies that elicited heart failure patients’ in-depth cognitive perceptions influencing their decision to participate in exercise and cardiac rehabilitation. Most previous research on cardiac rehabilitation utilization was focused on low referral rates and economic and environmental barriers to participation in cardiac rehabilitation^22^. While interventions focusing on increasing referrals and removing socio-economic barriers^23^ to cardiac rehabilitation participation are of a vital importance, based on our findings, just a simple increase in referrals to cardiac rehabilitation will likely not increase the rate of the uptake and completion of these programs if the patient-level cognitive barriers to exercise identified in the study are not addressed. Most of our participants with heart failure reported fear of exercise and low, or even absent, motivation to exercise. Interventions that will address these cognitive barriers can increase participation in cardiac rehabilitation.

Additionally, this is one of the first studies to oversample participants with HFpEF and investigate barriers and facilitators of participation in cardiac rehabilitation in HFpEF. While the evidence of cardiac rehabilitation benefits in HFpEF is still evolving, data from the REHAB-heart failure trial demonstrated that a tailored exercise program, implemented early in the course of HFpEF, improved physical functioning and quality of life^24^. Preliminary data suggest that exercise may be beneficial for HFpEF by reversing atrial remodeling and reducing diastolic dysfunction, or via peripheral mechanisms such as improving endothelial function in the skeletal muscle vasculature.^25^ Despite rising prevalence of HFpEF and worsening deleterious health outcomes, patients with HFpEF are underrepresented in randomized clinical trials (RCT) of cardiac rehabilitation to date.^19^ And, consequently, cardiac rehabilitation is not covered by a majority of health insurers for patients with HFpEF. Currently, one large RCT evaluating comprehensive physical rehabilitation for older adults with HFpEF is underway. In 2023, an AHA/ACC scientific statement on supervised exercise in HFpEF delineated a critical gap in knowledge regarding improving implementation of cardiac rehabilitation in HFpEF, specifically long-term adherence to cardiac rehabilitation and whether exercise should be augmented with other treatment modalities in such programs for HFpEF^26^. Our study findings contribute to bridging this significant gap in knowledge by demonstrating that weight management intervention should likely be combined with cardiac rehabilitation for HFpEF, as for some participants, weight was a barrier to exercise, while for others, the perceived benefit of weight loss was a motivator for participation in a supervised exercise program. Moreover, most of participants with HFPEF in our study expressed a desire to learn more and participate in a moderate and well supervised cardiac rehabilitation program. Including HFpEF as a qualifying condition for cardiac rehabilitation coverage by Medicare/Medicaid and private insurance plans can significantly increase physical activity level in this patient population and improved health outcomes.

Importantly, in our study, non-cardiac factors such as depression and chronic non-cardiac pain were important barriers to exercise among most of participants with both HFpEF and HFrEF. Depressive symptoms and perceived stress were high in our study sample. Prior studies also showed that depression is highly comorbid with heart failure: more than 20% of heart failure patients report depressive symptoms that are associated with poor health outcomes in heart failure.^27^ For HFpEF, depressive symptoms were associated with 1.5 increased risk for first heart failure hospitalization in the REGARDS study^28^. Clinical trials of pharmacological treatment for depression in heart failure have largely failed^29^. In contrast, in the heart failure-ACTION trial, HFrEF patients who were randomized to aerobic exercise achieved a 1.75-point reduction in depressive symptoms scores, compared to a 0.98-point reduction among heart failure patients who received usual care^4^. Corresponding to this RCT data, our study participants reported reduction in depressive symptoms as a perceived benefit of exercise and a motivating factor of adherence to the exercise program. Similar to depressive symptoms, chronic non-cardiac pain is a common and under-addressed symptom among patients with heart failure^30^. Improving chronic pain levels will likely encourage heart failure patients to adhere to a supervised exercise program.

This study has important clinical implications, as our findings provide a conceptual framework for patient-reported barriers and facilitators to participation in cardiac rehabilitation that can inform future intervention development focusing on uptake and adherence to cardiac rehabilitation among heart failure patients. Our study supports the notion that a supervised exercise program for heart failure will need to be supplemented with individual health psychologist counselling that uses motivational interviewing as a strategy to address cognitive processes behind patient decision-making regarding exercise. Heart failure patients may benefit from pre-cardiac rehabilitation or pre-referral counselling sessions that focus on addressing lack of information regarding cardiac rehabilitation programs, illness perceptions, fear of exercise and safety concerns, body image, low motivation, depressive symptoms, and chronic pain. For patients with HFpEF, one of the largest barriers to cardiac rehabilitation implementation is that the Centers for Medicare & Medicaid Services (CMS) currently do not include HFpEF as a covered qualifying condition for comprehensive cardiac rehabilitation. Our study offers, perhaps, a temporary but important solution to this problem by suggesting referrals to physical therapy.

HFpEF patients may also benefit from adding weight management to exercise programs. Finally, exploring alternative modes of cardiac rehabilitation delivery, such as hybrid and home-based cardiac rehabilitation may also increase intake of cardiac rehabilitation among heart failure patients^22^.

Limitations of the study include limited generalizability due to including heart failure patients from a single urban academic medical center. Patients with heart failure from rural areas, for example, may have other barriers to cardiac rehabilitation participation not reflected by this report. Strengths of this study include oversampling patients with HFpEF and AA patients from the Deep South, a geographical area disproportionately affected by heart failure morbidity and mortality. Additionally, this study used a robust approach to thematic analysis, an adequate sample size, and individual semi-structed interviews for data collection.

## Conclusion

In this diverse sample of adults with heart failure, mostly comprising of patients with HFpEF, levels of physical activity were low, but depression and perceived stress was high. Patients were not aware of cardiac rehabilitation programs and reported barriers to exercise that are modifiable by providing motivational counseling, advice, and referral from healthcare providers to existing cardiac rehabilitation programs. Patients reported on facilitators of cardiac rehabilitation participation that can inform future intervention development. Specifically, patients with HFpEF may benefit from referrals to existing and available-to-them programs, such as physical therapy, coupled with weight loss and depression management.

## Data Availability

All data produced in the present study are available upon reasonable request to the authors.

## Acknowledgments

Authors want to thank the patients with heart failure who provided their perspective for this report.

## Ethics approval and consent to participate

This study was approved by the University’s of Alabama at Birmingham IRB as an exempt protocol and consent to participate was waived by the IRB. Instead, all study participants were provided with a study information sheet that described study’s procedures, potential risks and benefits. This study adhered to the Declaration of Helsinki

## Consent for publication

Not applicable as no personal identifiable data or images are included in this manuscript.

## Author Contributions

YK: conceptualized the study design, supervised data collection, and drafted the initial manuscript. YK, MN: conducted analyses and contributed to interpretation of results. YK, MN: coordinated patient recruitment and managed study logistics. YK, MN, TB, AC, LH: reviewed and revised the manuscript for important intellectual content. All authors: approved the final version of the manuscript and agreed to be accountable for all aspects of the work.

---

**Supplemental Figure 1.**
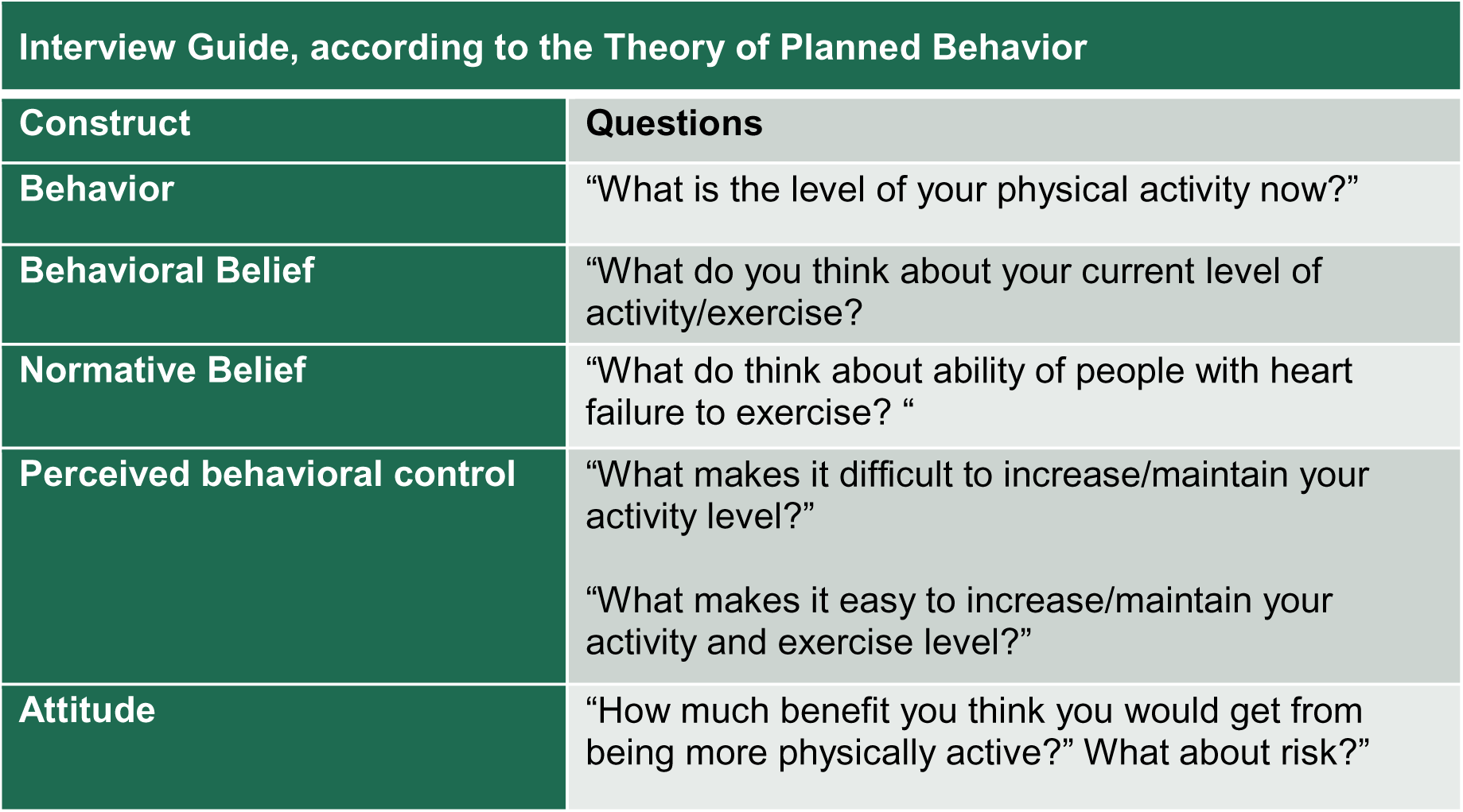

## REFERENCES

1. Virani SS, Alonso A, Benjamin EJ, et al. Heart Disease and Stroke Statistics-2020 Update: A Report From the American Heart Association. Circulation. Mar 3 2020;141(9):e139–e596. doi:10.1161/CIR.0000000000000757

2. Brown TM, Pack QR, Beregg EA, et al. Core Components of Cardiac Rehabilitation Programs: 2024 Update: A Scientific Statement From the American Heart Association and the American Association of Cardiovascular and Pulmonary Rehabilitation: Endorsed by the American College of Cardiology. J Cardiopulm Rehabil Prev. Mar 1 2025;45(2):E6–E25. doi:10.1097/Hcardiac rehabilitation.0000000000000930

3. Long L, Mordi IR, Bridges C, et al. Exercise-based cardiac rehabilitation for adults with heart failure. Cochrane Database Syst Rev. Jan 29 2019;1:CD003331. doi:10.1002/14651858.CD003331.pub5

4. Blumenthal JA, Babyak MA, O’Connor C, et al. Effects of exercise training on depressive symptoms in patients with chronic heart failure: the heart failure-ACTION randomized trial. JAMA. Aug 1 2012;308(5):465–74. doi:10.1001/jama.2012.8720

5. Scalvini S, Grossetti F, Paganoni AM, La Rovere MT, Pedretti RF, Frigerio M. Impact of in-hospital cardiac rehabilitation on mortality and readmissions in heart failure: A population study in Lombardy, Italy, from 2005 to 2012. Eur J Prev Cardiol. May 2019;26(8):808–817. doi:10.1177/2047487319833512

6. Wall HK, Stolp H, Wright JS, et al. The Million Hearts Initiative: CATALYZING UTILIZATION OF CARDIAC REHABILITATION AND ACCELERATING IMPLEMENTATION OF NEW CARE MODELS. Journal of Cardiopulmonary Rehabilitation and Prevention. 2020;40(5):290–293. doi:10.1097/hcr.0000000000000547

7. Keteyian SJ, Jackson SL, Chang A, et al. Tracking Cardiac Rehabilitation Utilization in Medicare Beneficiaries: 2017 UPDATE. Journal of Cardiopulmonary Rehabilitation and Prevention. 2022;42(4):235–245. doi:10.1097/hcr.0000000000000675

8. Brown TM, Zhang Y, McNamara M, et al. The American Association of Cardiovascular and Pulmonary Rehabilitation National Cardiac Rehabilitation Registry: Design and Participant Characteristics. J Cardiopulm Rehabil Prev. Jul 29 2025;doi:10.1097/Hcardiac rehabilitation.0000000000000976

9. Golwala H, Pandey A, Ju C, et al. Temporal Trends and Factors Associated With Cardiac Rehabilitation Referral Among Patients Hospitalized With Heart Failure: Findings From Get With The Guidelines-Heart Failure Registry. J Am Coll Cardiol. Aug 25 2015;66(8):917–26. doi:10.1016/j.jacc.2015.06.1089

10. Keshvani N, Subramanian V, Wrobel CA, et al. Patterns of Referral and Postdischarge Utilization of Cardiac Rehabilitation Among Patients Hospitalized With Heart Failure: An Analysis From the GWTG-heart failure Registry. Circ Heart Fail. Aug 2023;16(8):e010144. doi:10.1161/circheartfailure.122.010144

11. Ritchey MD, Maresh S, McNeely J, et al. Tracking Cardiac Rehabilitation Participation and Completion Among Medicare Beneficiaries to Inform the Efforts of a National Initiative. Circulation: Cardiovascular Quality and Outcomes. 2020;13(1):e005902. doi:doi:10.1161/CIRCOUTCOMES.119.005902

12. Bartholomew LK, Parcel GS, Kok G. Intervention mapping: a process for developing theory- and evidence-based health education programs. Health Educ Behav. Oct 1998;25(5):545–63. doi:10.1177/109019819802500502

13. Johnston DW, Johnston M, Pollard B, Kinmonth AL, Mant D. Motivation is not enough: prediction of risk behavior following diagnosis of coronary heart disease from the theory of planned behavior. Health Psychol. Sep 2004;23(5):533–8. doi:10.1037/0278-6133.23.5.533

14. Houle J, Gallani MC, Pettigrew M, et al. Acceptability of a computer-tailored and pedometer-based socio-cognitive intervention in a secondary coronary heart disease prevention program: A qualitative study. Digit Health. Jan-Dec 2020;6:2055207619899840. doi:10.1177/2055207619899840

15. Ajzen I, Brown TC, Carvajal F. Explaining the discrepancy between intentions and actions: the case of hypothetical bias in contingent valuation. Pers Soc Psychol Bull. Sep 2004;30(9):1108–21. doi:10.1177/0146167204264079

16. Wu JR, Lennie TA, Dunbar SB, Pressler SJ, Moser DK. Does the Theory of Planned Behavior Predict Dietary Sodium Intake in Patients With Heart Failure? West J Nurs Res. Apr 2017;39(4):568–581. doi:10.1177/0193945916672661

17. Beatty AL, Magnusson SL, Fortney JC, Sayre GG, Whooley MA. VA FitHeart, a Mobile App for Cardiac Rehabilitation: Usability Study. JMIR Hum Factors. Jan 15 2018;5(1):e3. doi:10.2196/humanfactors.8017

18. Austin Z, Sutton J. Qualitative research: getting started. Can J Hosp Pharm. Nov 2014;67(6):436–40. doi:10.4212/cjhp.v67i6.1406

19. Forman DE, Sanderson BK, Josephson RA, Raikhelkar J, Bittner V, American College of Cardiology’s Prevention of Cardiovascular Disease S. Heart Failure as a Newly Approved Diagnosis for Cardiac Rehabilitation: Challenges and Opportunities. J Am Coll Cardiol. Jun 23 2015;65(24):2652–2659. doi:10.1016/j.jacc.2015.04.052

20. Heidenreich PA, Bozkurt B, Aguilar D, et al. 2022 AHA/ACC/heart failureSA Guideline for the Management of Heart Failure: A Report of the American College of Cardiology/American Heart Association Joint Committee on Clinical Practice Guidelines. Circulation. May 3 2022;145(18):e895–e1032. doi:10.1161/CIR.0000000000001063

21. Heidenreich PA, Bozkurt B, Aguilar D, et al. 2022 AHA/ACC/heart failureSA Guideline for the Management of Heart Failure: Executive Summary: A Report of the American College of Cardiology/American Heart Association Joint Committee on Clinical Practice Guidelines. Circulation. May 3 2022;145(18):e876–e894. doi:10.1161/CIR.0000000000001062

22. Beatty AL, Beckie TM, Dodson J, et al. A New Era in Cardiac Rehabilitation Delivery: Research Gaps, Questions, Strategies, and Priorities. Circulation. Jan 17 2023;147(3):254–266. doi:10.1161/circulationaha.122.061046

23. Pandey A, Keshvani N, Zhong L, et al. Temporal Trends and Factors Associated With Cardiac Rehabilitation Participation Among Medicare Beneficiaries With Heart Failure. JACC: Heart Failure. 2021/07/01/ 2021;9(7):471–481. 10.1016/j.jchf.2021.02.006

24. Gilbert ON, Mentz RJ, Bertoni AG, et al. Relationship of Race With Functional and Clinical Outcomes With the REHAB-heart failure Multidomain Physical Rehabilitation Intervention for Older Patients With Acute Heart Failure. J Am Heart Assoc. Nov 7 2023;12(21):e030588. doi:10.1161/JAHA.123.030588

25. Keteyian SJ. Exercise training in patients with heart failure and preserved ejection fraction: findings awaiting discovery. J Am Coll Cardiol. Aug 13 2013;62(7):593–4. doi:10.1016/j.jacc.2013.01.098

26. Sachdev V, Sharma K, Keteyian SJ, et al. Supervised Exercise Training for Chronic Heart Failure With Preserved Ejection Fraction: A Scientific Statement From the American Heart Association and American College of Cardiology. Circulation. Apr 18 2023;147(16):e699–e715. doi:10.1161/cir.0000000000001122

27. Rutledge T, Reis VA, Linke SE, Greenberg BH, Mills PJ. Depression in heart failure a meta-analytic review of prevalence, intervention effects, and associations with clinical outcomes. J Am Coll Cardiol. Oct 17 2006;48(8):1527–37. doi:10.1016/j.jacc.2006.06.055

28. Khodneva Y, Goyal P, Levitan EB, et al. Depressive Symptoms and Incident Hospitalization for Heart Failure: Findings From the REGARDS Study. J Am Heart Assoc. Apr 5 2022;11(7):e022818. doi:10.1161/jaha.121.022818

29. Celano CM, Villegas AC, Albanese AM, Gaggin HK, Huffman JC. Depression and Anxiety in Heart Failure: A Review. Harv Rev Psychiatry. Jul/Aug 2018;26(4):175–184. doi:10.1097/HRP.0000000000000162

30. Vikan KK, Landmark T, Gjeilo KH. Prevalence of chronic pain and chronic widespread pain among subjects with heart failure in the general population: The HUNT study. Eur J Pain. Feb 2024;28(2):273–284. doi:10.1002/ejp.2176

